# Snakebite prevalence, burden, healthcare-seeking behavior, and climate change awareness: a cross-sectional household survey in rural Gulu District, northern Uganda

**DOI:** 10.64898/2026.09.14.26363011

**Authors:** Fauzi Ibrahim Ali, Asiimwe Lydia Irene, Kyeyune Henry, Swabrah Halimah Masaba, Isaac Ahamada Basule, Eva Laker Agnes Odongpiny, Stella Maris Nanyonga

**Affiliations:** College of Health Sciences, Makerere University, Kampala, Uganda; Directorate of Laboratory Services, National Drug Authority, Kampala, Uganda; Infectious Diseases Institute, Makerere University, Kampala, Uganda; Medicine Quality Research Group (MQRG), NDM Centre for Global Health Research, Nuffield Department of Medicine, University of Oxford, Oxford, United Kingdom

## Abstract

**Background:** Snakebite envenoming (SBE) is a WHO-listed neglected tropical disease affecting mainly rural communities in sub-Saharan Africa. Northern Uganda is a recognized high-burden area, but surveillance depends on health facility records, which cannot count victims who never reach care. Community-level data are therefore scarce, and awareness of climate change as a potential driver is unknown. We determined the household prevalence, health outcomes, socioeconomic burden, knowledge, attitudes, and climate change awareness of SBE in rural Gulu District.

**Methodology/Principal findings:** We conducted a cross-sectional household survey in Palaro Subcounty, Gulu District, using multi-stage cluster sampling. A structured questionnaire was administered to 244 consenting household heads or knowledgeable adults; estimates were design-weighted. Lifetime household prevalence was 36.9% (95% CI 20.0-57.7). Most bites occurred while walking or farming, on roads or in fields, and at night, peaking in May. Among the most recent events, 7/90 (7.8%) ended in death and 4 (4.4%) in permanent disability. Five of the seven deaths occurred before the victim reached a health facility. A traditional healer was the first source of care for 50.6% of victims, whereas only 2.2% presented first to a hospital. The median estimated cost per event (78,333 UGX) was about seven-tenths of mean monthly household income, yet the median direct cost was zero: the loss was almost entirely forgone labor (median 14 days lost). Awareness of climate change was high (91.5%), and 59.7% believed it affects snakebite.

**Conclusions/Significance:** More than a third of households have been affected by snakebite. Still, most of this burden is invisible to facility-based surveillance: deaths occur before arrival, first care is sought outside the formal system, and economic loss runs through forgone labor rather than medical fees. Antivenom access alone will be insufficient unless victims reach care sooner. High community awareness of climate change offers an entry point for integrated prevention.

**Author summary:** Snakebite is a neglected tropical disease that kills or disables mainly poor farming families. In much of Africa, we do not know how common it truly is, because most victims never reach a hospital. We visited 258 households in rural Gulu District, northern Uganda, and 244 agreed to take part. We asked about any snakebites the household had experienced, what happened during the most recent one, what it cost, and what people knew and believed about snakebite and their environment. More than a third of households had experienced a snakebite. Most bites happened while walking or farming, on roads or in fields, and at night, with a clear peak in May. Roughly one in thirteen victims died, and most of those deaths happened before the person reached a health facility. Half of victims went first to a traditional healer rather than to a clinic, and an episode cost close to a family’s whole monthly income. Larger and better-off households were more likely to be affected. Almost everyone had heard of climate change, and most thought it affected snakebite. Our results show a heavy, under-recognized burden and point to pre-hospital care, antivenom access, and community education as priorities.

## Introduction

Snakebite envenoming (SBE) is a World Health Organization-listed neglected tropical disease (NTD) that causes an estimated 1.8-2.7 million envenomings and up to 138,000 deaths each year, with a further 400,000 survivors left with permanent physical or psychological sequelae [1,2]. The burden falls overwhelmingly on rural agricultural communities in low- and middle-income countries, where high occupational exposure converges with limited access to effective care [1,3,4].

In sub-Saharan Africa, SBE is a major but poorly measured public health problem, with approximately 435,000-580,000 bites and around 32,000 deaths annually, the great majority occurring in rural settings [3,5,6]. Facility-based surveillance substantially underestimates the true incidence because a large share of cases, up to 70%, never reach the formal health system, owing to reliance on traditional healers, financial barriers, and long travel distances [3,5,7]. Uganda, and northern Uganda in particular, is a recognized high-burden area [8,9]. Gulu District combines a tropical climate, medically important snake species, and an agrarian economy that heighten human-snake contact, with community incidence estimated at around 101 cases per 100,000 population [9,10,11]. Even when victims reach healthcare facilities, outcomes are compromised by inadequate provider knowledge, inconsistent antivenom supply, absent treatment protocols, and weak referral systems, while cultural preference for traditional healers may further delay access to effective treatment [8,9,12].

An emerging concern is the influence of climate change on snakebite risk. Climatic conditions shape the distribution, activity, and habitat suitability of venomous snakes, while land-use changes, mainly agricultural expansion, create ecotones that can intensify human-snake contact [13,14,15]. Predictive models suggest range shifts for venomous species in East Africa under future climate scenarios [14,15]. Whether rural communities perceive this potential driver has not, to our knowledge, been reported in northern Uganda, representing a critical knowledge gap and a missed opportunity for integrated prevention strategies that combine public health interventions and environmental education.

Three critical evidence gaps persist in Uganda: limited community-level prevalence data, particularly in Gulu District; a lack of quantitative evidence on the socioeconomic burden of SBE; and scarce local evidence on ecological and climatic drivers of snakebite risk. To address these gaps, we conducted a cross-sectional household survey in rural Gulu District to estimate the household prevalence and health outcomes of SBE; characterize its individual- and community-level burden and associated healthcare-seeking behaviors; and assess community knowledge, attitudes, and awareness of SBE, including perceptions of climate change as a potential driver of snakebite incidence.

## Methods

### Ethics statement

Written informed consent was obtained from every participant before enrolment, after the study purpose, procedures, and voluntary nature were explained in the participant’s preferred language (English or Luo/Acholi). Participants were free to decline or withdraw without consequence, and data were de-identified and kept confidential. The study was approved by the Makerere University School of Health Sciences Research and Ethics Committee (MAKSHSREC-2025-646) and registered with the Uganda National Council for Science and Technology (HS7325ES).

### Study design and setting

This paper reports the quantitative component of a broader embedded mixed-methods study. The quantitative component was a cross-sectional household survey conducted in Palaro Subcounty, Aswa County, Gulu District, northern Uganda. Palaro is a rural, agriculture-dependent subcounty with six parishes (Awich, Labworomor, Mede, Ocetoaka, Oroko and Ongedo). According to the 2024 national census, it has a household population of 10,741 residents across 2,447 households and a mean household size of 4.4 [16]. Palaro lies approximately 50 kilometers from the nearest referral hospital, St. Mary’s Lacor Hospital. Data were collected between 20 and 24 April 2026.

### Study population and sampling

The study population comprised households of permanent residents of the sampled villages; the respondent was the household head or another adult with comprehensive knowledge of the household’s health history. A multi-stage cluster sampling strategy was used. In the first stage, systematic sampling with probability proportional to size (PPS) applied to the 2024 household census (2,447 households; k = 815.67; r = 815) selected three of six parishes: Labworomor, Ocetoaka and Ongedo. In the second stage, all 13 villages within the selected parishes were enumerated as clusters; because enumeration was complete, this stage contributed no additional selection probability. In the third stage, households were systematically sampled within each village using local council demographic records.

### Sample size

The target sample of 261 households was derived from the Cochran formula for a single proportion [17,18]. We assumed a household prevalence of 10%, a 5% margin of error, an intracluster correlation of 0.05, and a mean cluster size of 15, giving a design effect of 1.7, and applied a 10% non-response allowance. Realized enrolment is reported at the start of the Results. Reporting follows the STROBE statement for cross-sectional studies (S1 File).

### Data collection and variables

Trained research assistants fluent in English and Luo administered a structured questionnaire (S2 File), adapted from a validated household survey and contextualized to Gulu, through face-to-face interviews [19]. The instrument captured socio-demographic and socioeconomic characteristics (Section A); snakebite experience and the characteristics and outcomes of the most recent event (Section B); healthcare-seeking behaviours and the direct and indirect socioeconomic burden (Section C); and knowledge, attitudes, awareness and climate change perceptions (Section D). The principal outcome was household lifetime snakebite experience. Period prevalences were defined as a household respondent reporting at least one bite within the past five years and past twelve months, respectively. Monthly household income and treatment cost were collected in bands. To ensure accuracy, two individuals independently entered the data and cross-checked entries for discrepancies. We then cleaned the data in Microsoft Excel, applying range and skip-pattern checks, before analysis.

### Statistical analysis

All analyses were performed in Stata 15.1, with statistical significance set at two-sided α = 0.05. Item-level missingness was low (≤4% for most variables; highest for monthly income at 10.7%). We therefore performed complete-case analyses with variable-specific denominators; no missing values were imputed. The survey-weighted regression is exploratory and hypothesis-generating (p-values are not adjusted for multiplicity). Categorization was fixed before modelling: age (18-29, 30-44, 45-59, ≥60 years), household size (1-3, 4-6, ≥7 persons), education (none, primary, secondary, tertiary) and occupation (farmer vs non-farmer). For the burden analysis, band midpoints were imputed (open-ended bands valued at 1.3× the lower bound). Indirect costs were valued using a human-capital approach. Because subsistence agriculture lacks a formal working week, we treated all calendar days as productive and imputed daily income as the midpoint of the monthly household-income band divided by 30. Multiplying this daily rate by respondent-reported days lost yielded individual indirect costs, which we summed to obtain a total indirect cost and also expressed as a multiple of monthly household income. This deliberately avoids the upward wage bias of a formal-sector working calendar; applying alternative day-count conventions to the reported days gives broadly similar aggregate totals.

To account for the multi-stage design, we declared the analysis as a survey in Stata, specifying parish as the stratum and village as the primary sampling unit (svyset village [pweight=weight], strata(parish)). Because only three of six parishes were sampled, treating parish as a stratum is a certainty or self-representing approximation that omits the between-parish variance component and is therefore anti-conservative; we adopted it because three primary sampling units left too few degrees of freedom for stable variance estimation, and we flag the resulting design-based estimates accordingly. Each household was assigned a sampling weight equal to the number of households in the source population that it represented. Because selection occurred in two probability-bearing stages, this weight was the product of two components: the first accounted for the PPS selection of parishes, and the second for the systematic selection of households within each fully enumerated village. Multiplying the two components produced the final household weight, correcting for unequal probabilities of selection at both stages so that estimates represent the study population rather than only the households interviewed.

Continuous variables were summarized as mean (SD) or median (IQR) as appropriate, and categorical variables as frequencies and percentages of non-missing responses. Lifetime, five-year, and twelve-month prevalence were estimated as survey-adjusted proportions with 95% confidence intervals, and the design effect (DEFF) was reported. Bitten and non-bitten households were compared using the chi-square test (or Fisher’s exact test) and the t-test (or Mann-Whitney U test), as appropriate.

Factors independently associated with snakebite were examined using survey-weighted logistic regression with linearized (Taylor-series) variance estimation; crude odds ratios (OR) came from single-predictor models and adjusted odds ratios (aOR) from a multivariable model containing sex, age group, education, occupation (farmer vs non-farmer), income band and household-size category, each compared with its reference (lowest or first) category and all entered simultaneously, with 95% confidence intervals (CI). Several categories were sparse (12 non-farmers, five tertiary-educated respondents and a single household in the highest income band, all bitten), so estimates for these cells reflect near-separation and are unstable. With only 10 design degrees of freedom (13 villages minus 3 strata), the multivariable model is over-parameterized, and its estimates are reported as exploratory.

No formal subgroup or interaction analyses were prespecified or fitted. Differences in climate change awareness and in the belief that climate change affects snakebite were examined descriptively across education, age group, sex, and personal snakebite experience using Fisher’s exact test; these comparisons are unadjusted and exploratory, and no interaction terms were included in the regression models.

## Results

### Study population

Of the 261 households sampled, 3 were unreachable. Of the remaining 258 approached, 244 (94.6% of those approached) consented and were included in the final analysis; 90 (36.9%) reported ever having a household member bitten by a snake (Fig 1).

**Fig 1.**
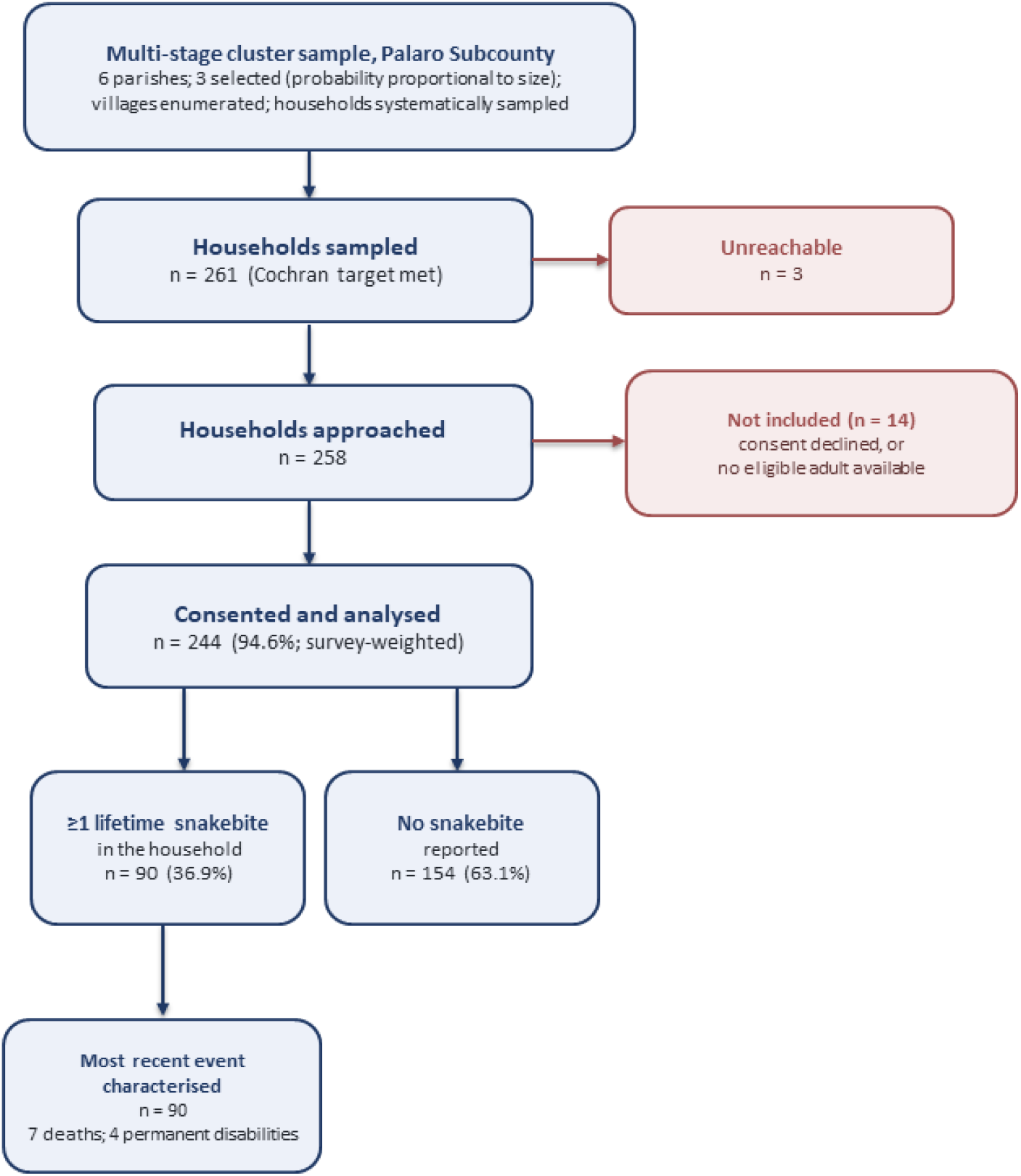
Participant flow and analytic denominators. Households sampled, approached, consenting and analyzed, with reasons for non-inclusion and the split by lifetime snakebite status.

### Participant characteristics

Respondents were evenly split by sex (50.6% male), with a mean age of 45.6 years (SD 13.2); 95% reported being farmers, and 72.9% reported a monthly income below 100,000 UGX. Households with and without reported snakebite experience were similar in age, sex, marital status, and household size, but differed significantly in imputed monthly income (164,198 vs 79,562 UGX; p < 0.001), education (p = 0.016), occupation (p = 0.010), and income band (p < 0.001) (Table 1). Denominators vary by item because of missingness (range 218-244); percentages use non-missing denominators.

**Table 1.** Baseline characteristics of households by snakebite status.

| Characteristic | Overall | Ever bitten | Not bitten | Test | *p* |
| --- | --- | --- | --- | --- | --- |
| Age of respondent, years - mean (SD) | 45.6 (13.2) | 47.4 (13.0) | 44.7 (13.3) | t-test | .139 |
| Household size - mean (SD) | 6.8 (3.0) | 7.0 (2.4) | 6.6 (3.3) | Mann-Whitney | .095 |
| Imputed monthly income, UGX - mean (SD) | 111,009 (114,674) | 164,198 (149,632) | 79,562 (71,629) | Mann-Whitney | <.001 |
| Age group, years - n (%) |  |  |  |  |  |
| 18-29 | 31 (13.2) | 8 (9.5) | 23 (15.3) | Fisher exact | .611 |
| 30-44 | 84 (35.9) | 30 (35.7) | 54 (36.0) |  |  |
| 45-59 | 82 (35.0) | 31 (36.9) | 51 (34.0) |  |  |
| 60+ | 37 (15.8) | 15 (17.9) | 22 (14.7) |  |  |
| Sex - n (%) |  |  |  |  |  |
| Male | 122 (50.6) | 48 (53.9) | 74 (48.7) | Fisher exact | .505 |
| Female | 119 (49.4) | 41 (46.1) | 78 (51.3) |  |  |
| Education - n (%) |  |  |  |  |  |
| None | 34 (14.3) | 16 (18.4) | 18 (11.9) | Fisher exact | .016 |
| Primary | 171 (71.8) | 53 (60.9) | 118 (78.1) |  |  |
| Secondary | 28 (11.8) | 14 (16.1) | 14 (9.3) |  |  |
| Tertiary | 5 (2.1) | 4 (4.6) | 1 (0.7) |  |  |

| Marital Status - n (%) |  |  |  |  |  |
| --- | --- | --- | --- | --- | --- |
| Single | 18 (7.5) | 6 (6.7) | 12 (7.9) | Fisher exact | .873 |
| Married | 177 (73.8) | 64 (71.9) | 113 (74.8) |  |  |
| Widowed | 36 (15.0) | 15 (16.9) | 21 (13.9) |  |  |
| Divorced | 9 (3.8) | 4 (4.5) | 5 (3.3) |  |  |
| Occupation - n (%) |  |  |  |  |  |
| Non-farmer | 12 (5.0) | 9 (10.2) | 3 (2.0) | Fisher exact | .010 |
| Farmer | 226 (95.0) | 79 (89.8) | 147 (98.0) |  |  |
| Monthly income band - n (%) |  |  |  |  |  |
| <100,000 UGX | 159 (72.9) | 45 (55.6) | 114 (83.2) | Fisher exact | <.001 |
| 100,000-300,000 | 38 (17.4) | 18 (22.2) | 20 (14.9) |  |  |
| 301,000-500,000 | 20 (9.2) | 17 (21.0) | 3 (2.2) |  |  |
| >500,000 | 1 (0.5) | 1 (1.2) | 0 (0.0) |  |  |

Comparison tests are unweighted (Fisher's exact test for categorical variables; t-test or Mann-Whitney U test for continuous variables). Denominators vary by item because of missingness (range 218-244); percentages use non-missing denominators. UGX, Ugandan shillings; SD, standard deviation.

### Prevalence and health outcomes

Survey-weighted household prevalence of snakebite was 36.9% (95% CI 20.0-57.7) over the lifetime, 29.1% (16.7-45.8) over five years, and 22.3% (10.2-42.0) over twelve months (Table 2). Among the 90 households reporting a bite, the most recent event was reported to occur while walking in 49 (54.4%), on a road in 42 (46.7%), or in a field in 38 (42.2%), and at night in 42 of 88 (47.7%). Seven of the 90 most recent events (7.8%) ended in death and four (4.4%) in permanent disability; five of the seven deaths occurred before the victim reached a health facility. Reported events clustered in May (Fig 2).

**Table 2.** Snakebite prevalence and health outcomes.

| Indicator | n/N | Estimate | 95% CI |
| --- | --- | --- | --- |
| Lifetime snakebite (household) | 90/244 | 36.9% | 20.0-57.7 |
| Snakebite in past 5 years (household) | 72/244 | 29.1% | 16.7-45.8 |
| Snakebite in the past 12 months (household) | 52/244 | 22.3% | 10.2-42.0 |
| **Victim's sex (most recent case)** |  |  |  |
| Male | 46/90 | 51.1% |  |
| Female | 44/90 | 48.9% |  |
| **Activity at time of bite** |  |  |  |
| Farming | 31/90 | 34.4% |  |
| Walking | 49/90 | 54.4% |  |
| Sleeping | 4/90 | 4.4% |  |
| Fetching water | 5/90 | 5.5% |  |
| Other | 1/90 | 1.1% |  |
| **Place of bite** |  |  |  |
| Home/Indoors | 7/90 | 7.8% |  |
| Field | 38/90 | 42.2% |  |
| Road | 42/90 | 46.7% |  |
| Other | 3/90 | 3.3% |  |
| **Time of bite** |  |  |  |
| Morning | 30/88 | 34.1% |  |
| Afternoon | 16/88 | 18.2% |  |
| Night | 42/88 | 47.7% |  |
| **Outcome of the bite** |  |  |  |
| Full recovery | 79/90 | 87.8% |  |
| Permanent disability | 4/90 | 4.4% |  |
| Death | 7/90 | 7.8% |  |
| **Death occurred before reaching the facility.** |  |  |  |
| Yes | 5/7 | 71.4% |  |
| No | 2/7 | 28.6% |  |
| **Victim age, median (IQR)** |  | 26 (18-34) |  |

**Fig 2.**
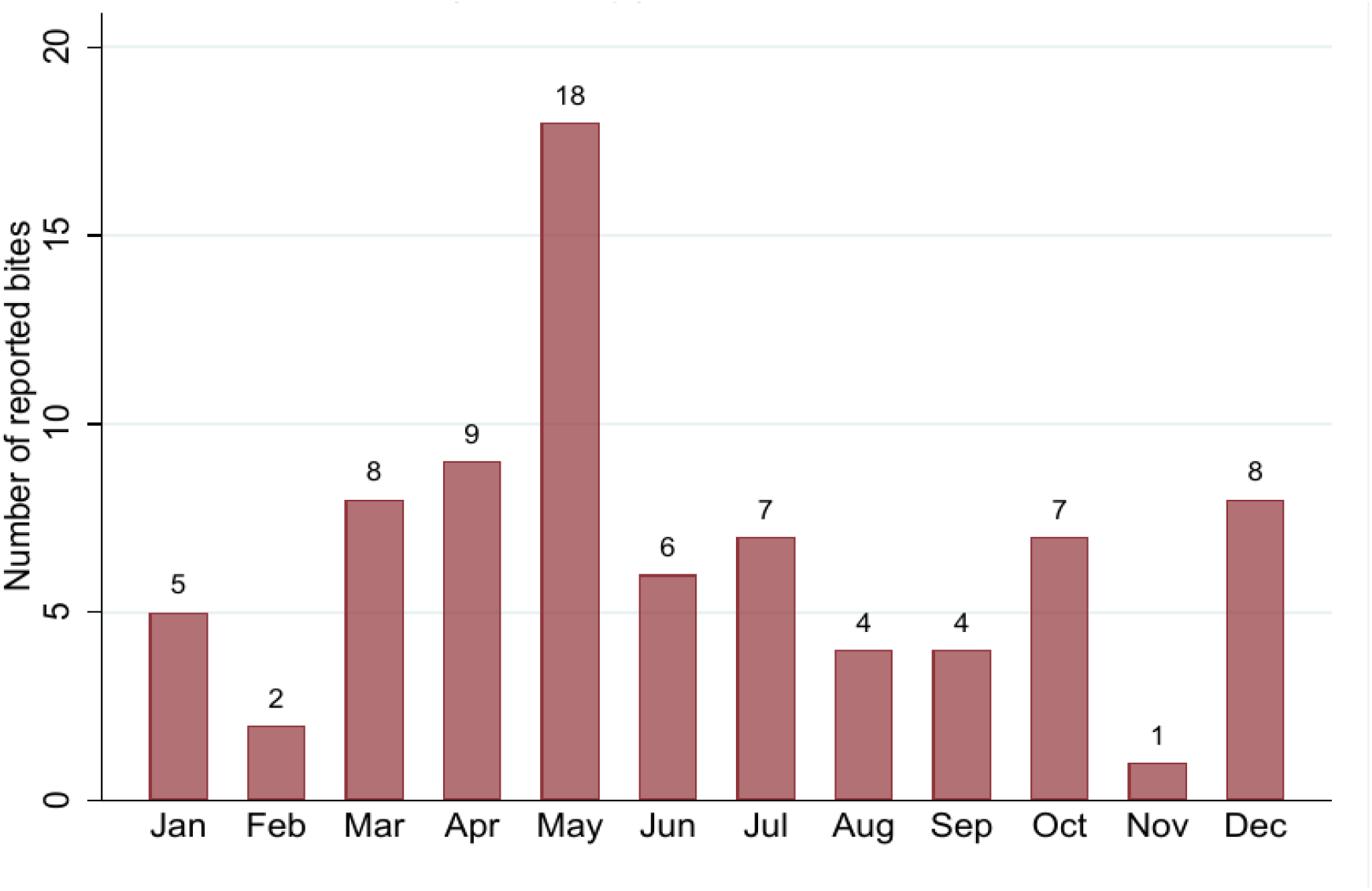
Monthly distribution of reported snakebite events. Counts refer to the most recent reported event in each affected household for which the month was recorded (n = 79).

### Design effect

The realised design effects (DEFF 5.26-8.20; DEFT 2.29-2.86) reduced the effective sample size for the lifetime estimate from 244 to approximately 30, a loss of about 88%. This implies an intracluster correlation of roughly 0.24-0.41, far above the 0.05 assumed at design (Table 3). The weighted and unweighted lifetime point estimates were identical (36.9%; unweighted 95% CI 31.0-43.1), consistent with an approximately self-weighting design; weighting affected the variance rather than the point estimate.

**Table 3.** Design effect and weighted versus unweighted sensitivity analysis.

| Prevalence measure | Unweighted % (95% CI) | Weighted % (95% CI) | DEFF | DEFT |
| --- | --- | --- | --- | --- |
| Lifetime | 36.9 (31.0-43.1) | 36.9 (20.0-57.7) | 8.20 | 2.86 |
| Past 5 years | 29.5 (24.1-35.6) | 29.1 (16.7-45.8) | 5.26 | 2.29 |
| Past 12 months | 21.3 (16.6-26.9) | 22.3 (10.2-42.0) | 7.26 | 2.69 |

DEFF, design effect; DEFT, design factor ( $DEFT = \sqrt{DEFF}$ ). $DEFF > 1$ indicates variance inflation from clustering.

### Healthcare-seeking behavior and socioeconomic burden

A traditional healer was reported to be the first source of care for half of victims (50.6%), with 37.1% first attending a health center, and 2.2% a hospital (Table 4). Although 68.5% reported having reached some form of care within an hour, the median distance to a facility was 4 km (IQR 3-8). Nearly half (47.7%) reported that care was free, but among those who reported paying, relatives most often covered the cost (79.1%). A median of 14 work or school days (IQR 7-21) was reported lost per event. The median total estimated economic cost per event was 78,333 UGX (IQR 41,667-160,000), approximately seven-tenths of the mean monthly household income (Table 4); because the median direct cost was zero, this total is driven almost entirely by imputed indirect (productivity) cost.

**Table 4.**
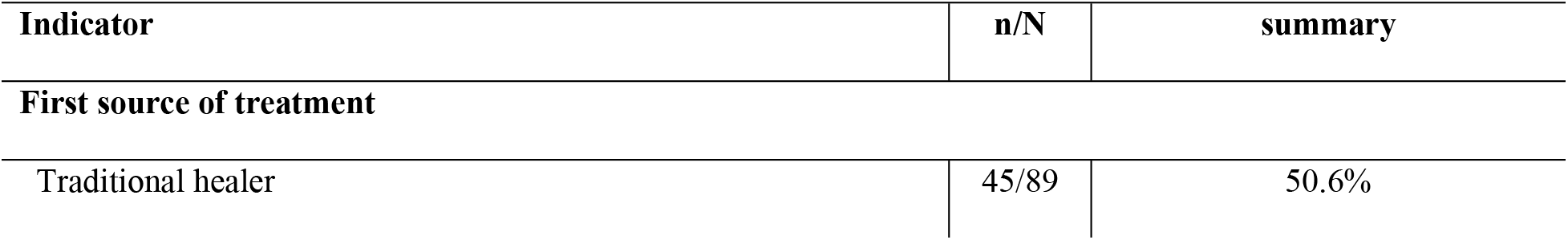
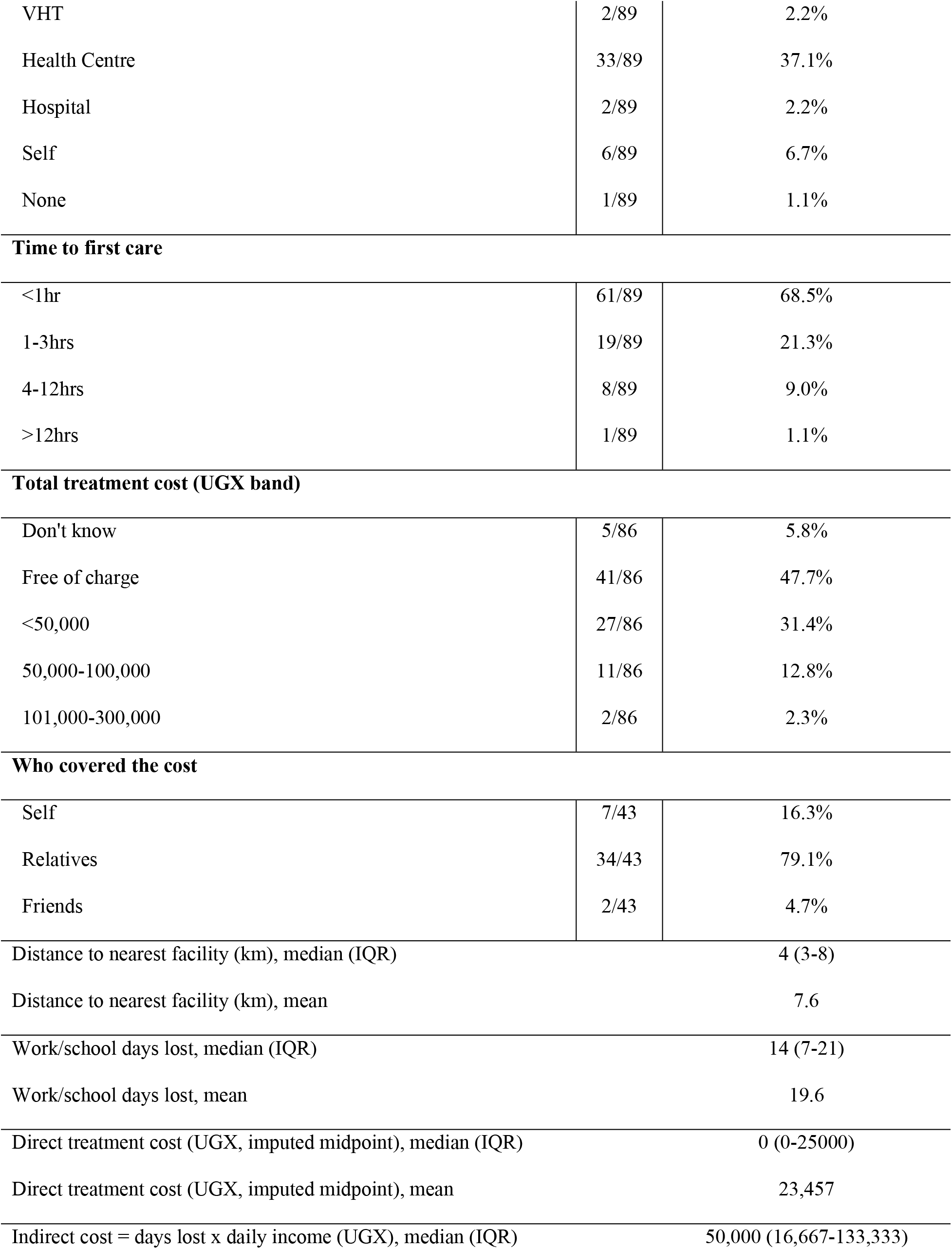
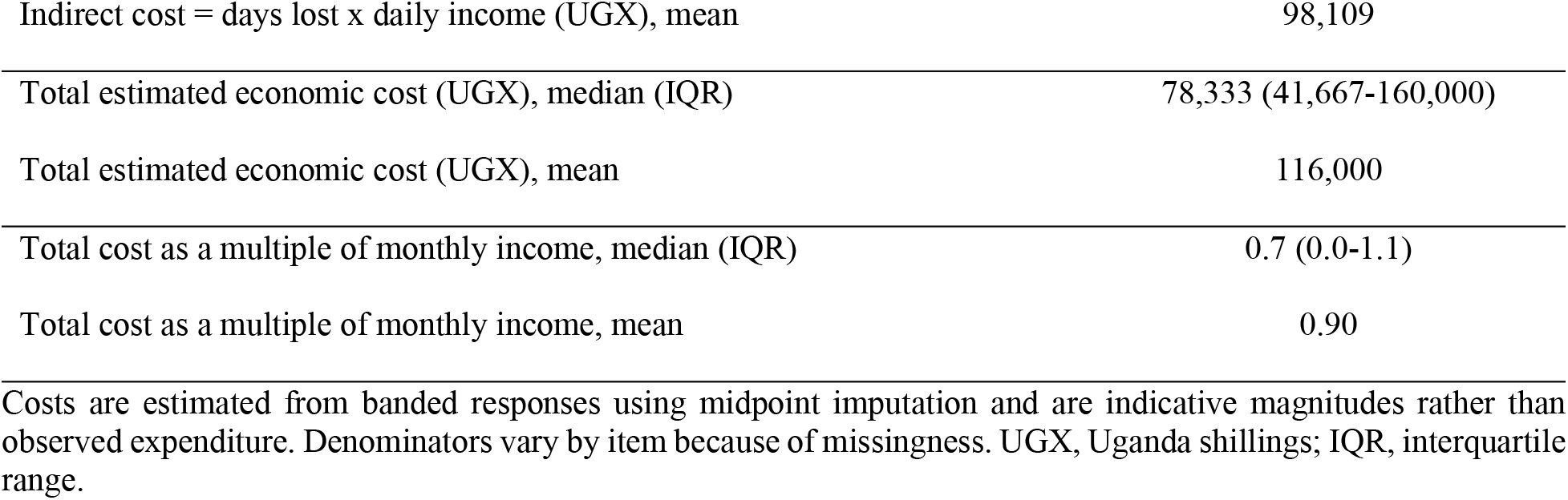
Healthcare-seeking behavior and socioeconomic burden among affected households.

### Knowledge, attitudes and awareness

About two-thirds of respondents (68.3%) reported having heard of snakebite envenoming. While 89.2% reported believing that hospital treatment is effective, 66.5% also reported believing that traditional treatment is effective. Reported preferred first care was divided between a health facility alone (33.5%) and both traditional and formal care (37.6%). As for perceived risk factors, bushes (63.0%) and night movement (28.4%) were most frequently reported, whereas the most commonly cited preventive measures were wearing boots (66.3%) and bush clearing (40.3%) (Table 5).

**Table 5.**
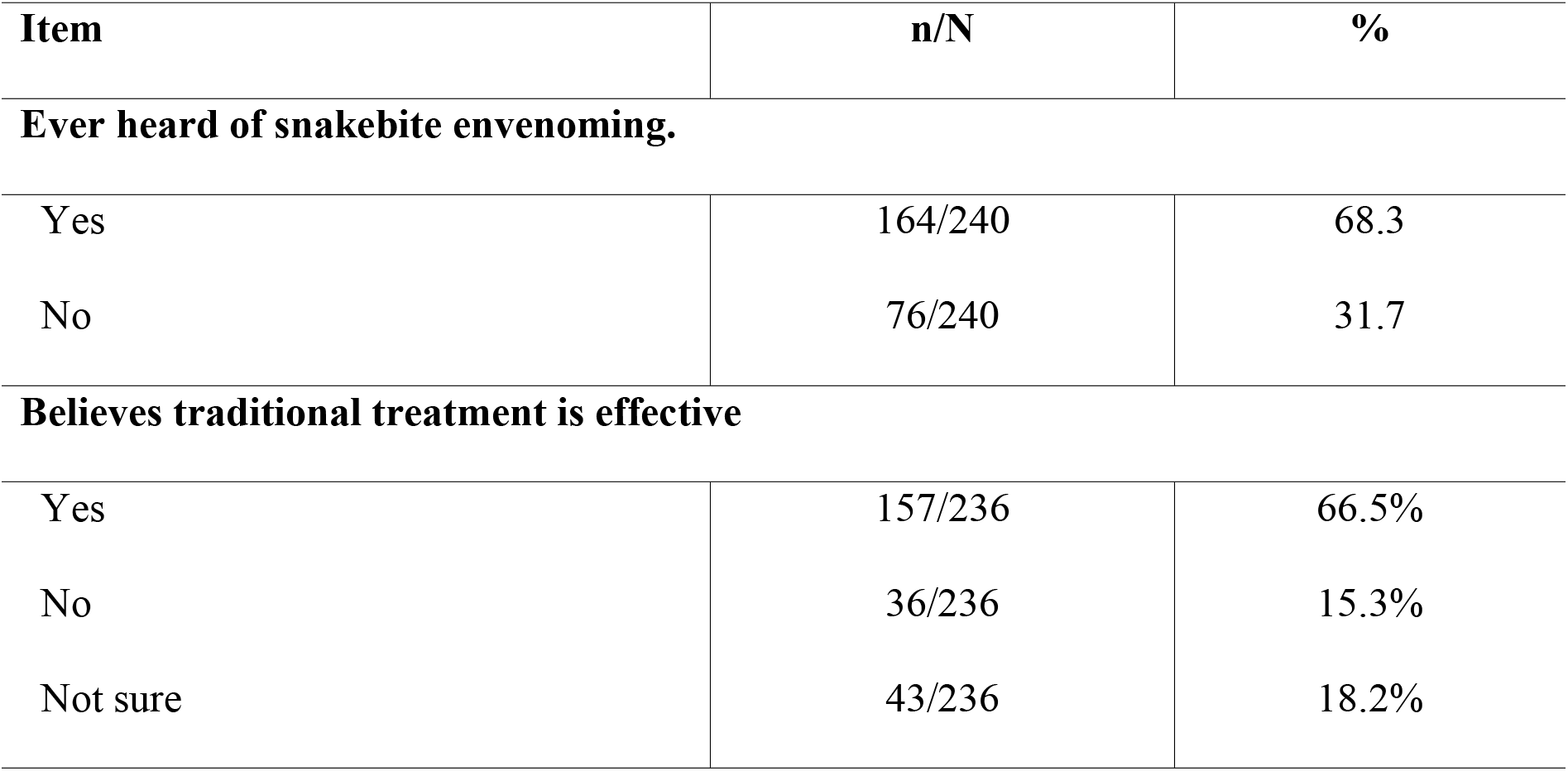
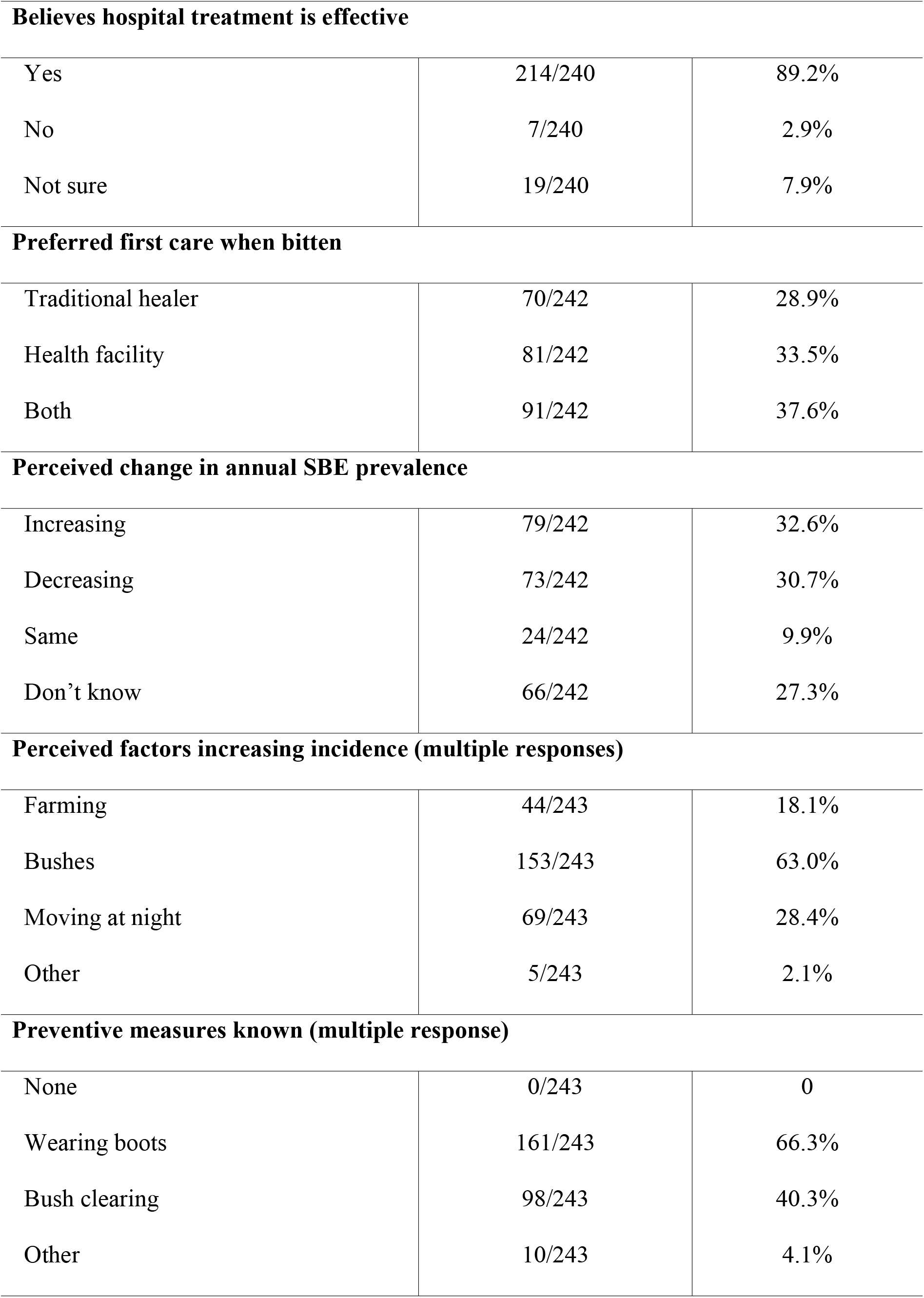
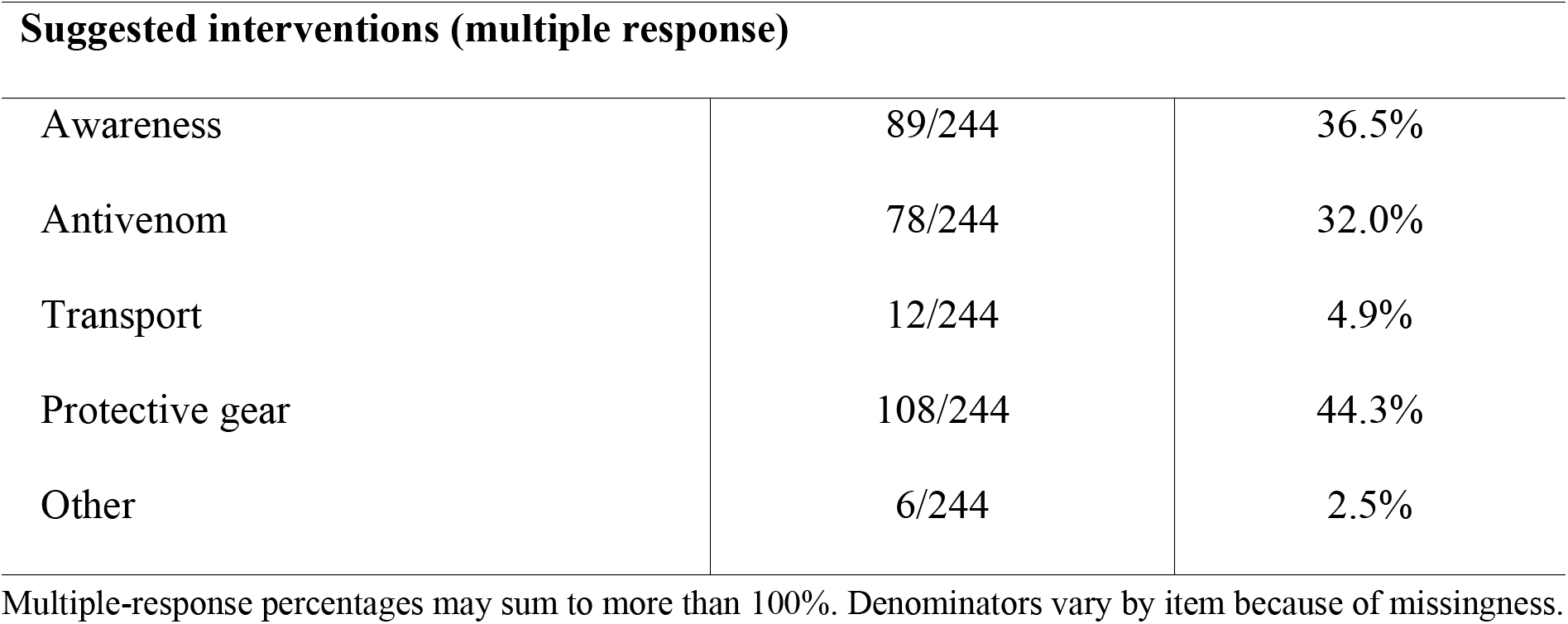
Knowledge, attitudes and awareness of snakebite.

| Item | n/N | % |
| --- | --- | --- |
| **Ever heard of snakebite envenoming.** |  |  |
| Yes | 164/240 | 68.3 |
| No | 76/240 | 31.7 |
| **Believes traditional treatment is effective** |  |  |
| Yes | 157/236 | 66.5% |
| No | 36/236 | 15.3% |
| Not sure | 43/236 | 18.2% |
| **Believes hospital treatment is effective** |  |  |
| Yes | 214/240 | 89.2% |
| No | 7/240 | 2.9% |
| Not sure | 19/240 | 7.9% |
| **Preferred first care when bitten** |  |  |
| Traditional healer | 70/242 | 28.9% |
| Health facility | 81/242 | 33.5% |
| Both | 91/242 | 37.6% |
| **Perceived change in annual SBE prevalence** |  |  |
| Increasing | 79/242 | 32.6% |
| Decreasing | 73/242 | 30.7% |
| Same | 24/242 | 9.9% |
| Don't know | 66/242 | 27.3% |
| **Perceived factors increasing incidence (multiple responses)** |  |  |
| Farming | 44/243 | 18.1% |
| Bushes | 153/243 | 63.0% |
| Moving at night | 69/243 | 28.4% |
| Other | 5/243 | 2.1% |
| **Preventive measures known (multiple response)** |  |  |
| None | 0/243 | 0 |
| Wearing boots | 161/243 | 66.3% |
| Bush clearing | 98/243 | 40.3% |
| Other | 10/243 | 4.1% |
| Suggested interventions (multiple response) |  |  |
| Awareness | 89/244 | 36.5% |
| Antivenom | 78/244 | 32.0% |
| Transport | 12/244 | 4.9% |
| Protective gear | 108/244 | 44.3% |
| Other | 6/244 | 2.5% |

Multiple-response percentages may sum to more than 100%. Denominators vary by item because of missingness.

### Factors associated with snakebite experience

In survey-weighted logistic regression, higher household income was strongly and independently associated with snakebite experience: households in the 301,000-500,000 UGX band had over ten times the odds of those in the lowest band (aOR 10.76, 95% CI 5.35-21.62; p < 0.001). Larger household size was also independently associated (4-6 persons: aOR 6.26, 95% CI 1.26-31.19; ≥7 persons: aOR 13.17, 95% CI 1.44-120.26), although with wide confidence intervals reflecting small strata. Sex, age group, and education were not independently associated after adjustment, and a crude protective association for farmers did not persist (Table 6).

**Table 6.**
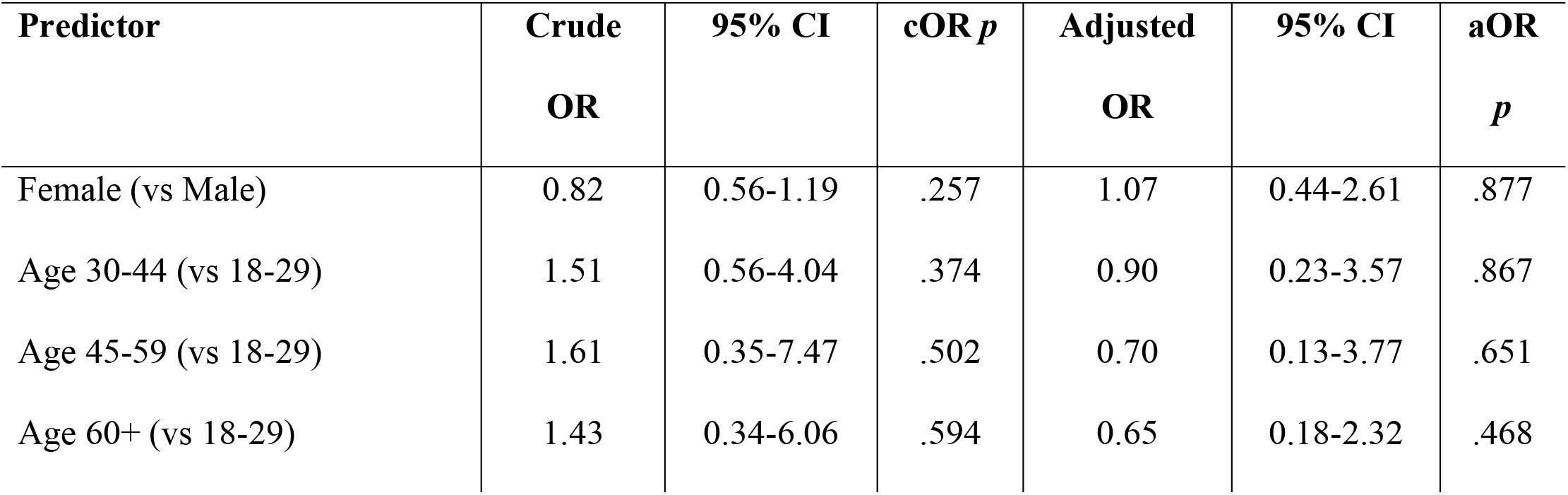
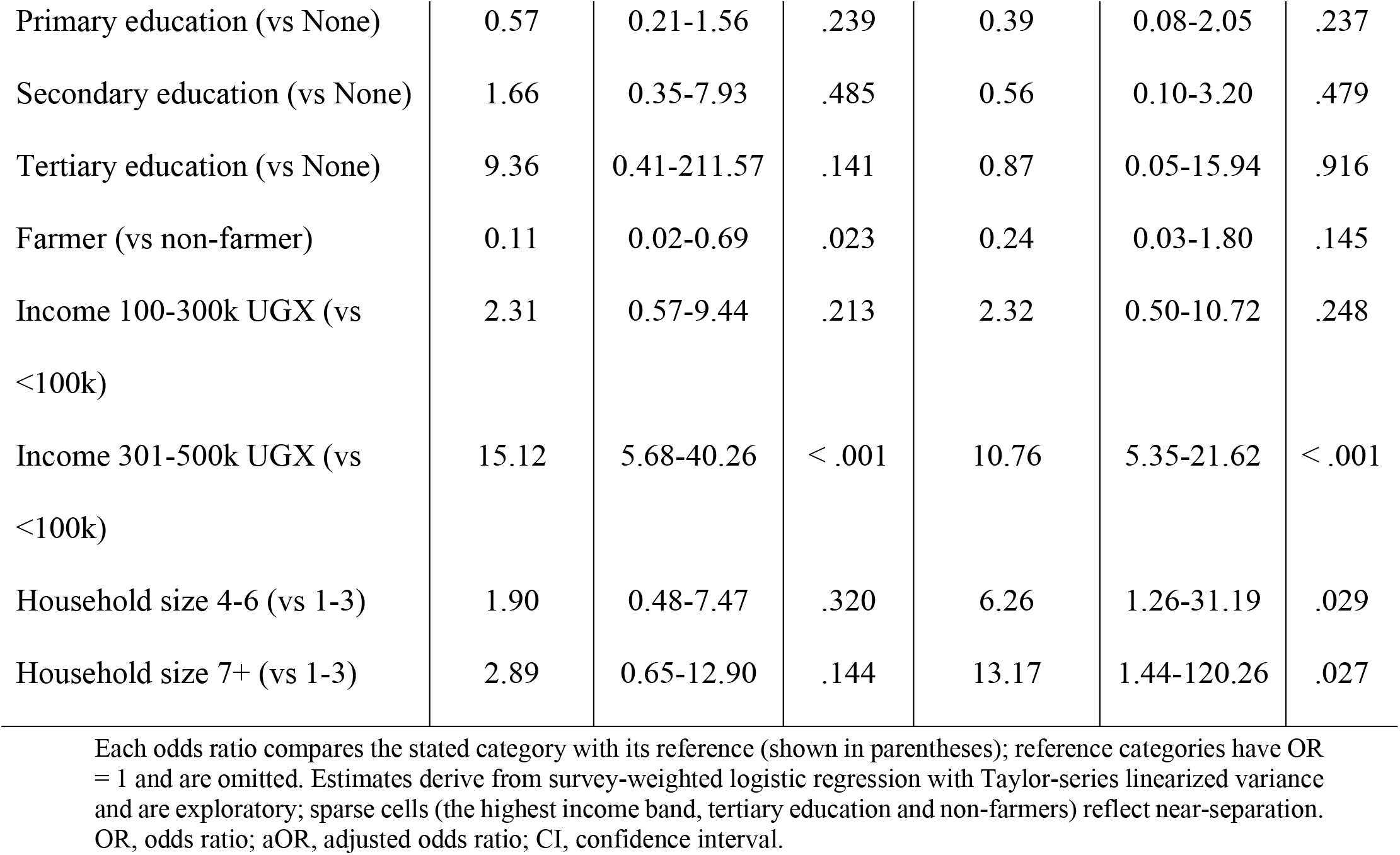
Factors associated with household snakebite experience.

### Climate change awareness and perceptions

Climate change awareness was high, with 91.5% reporting having heard about it. Regarding its link to snakebite, 59.7% reported believing that such a link exists, while a quarter (25.4%) reported being unsure. The most commonly reported environmental changes were higher temperatures (43.6%) and deforestation (38.7%); reasons cited for a perceived change in snakebite trends were land use (49.7%) and weather (37.3%). Having heard of climate change increased with education (Fisher’s exact p = 0.010), whereas the belief that climate change affects snakebite did not differ significantly by education, age, sex, or personal snakebite experience (Table 7).

**Table 7.**
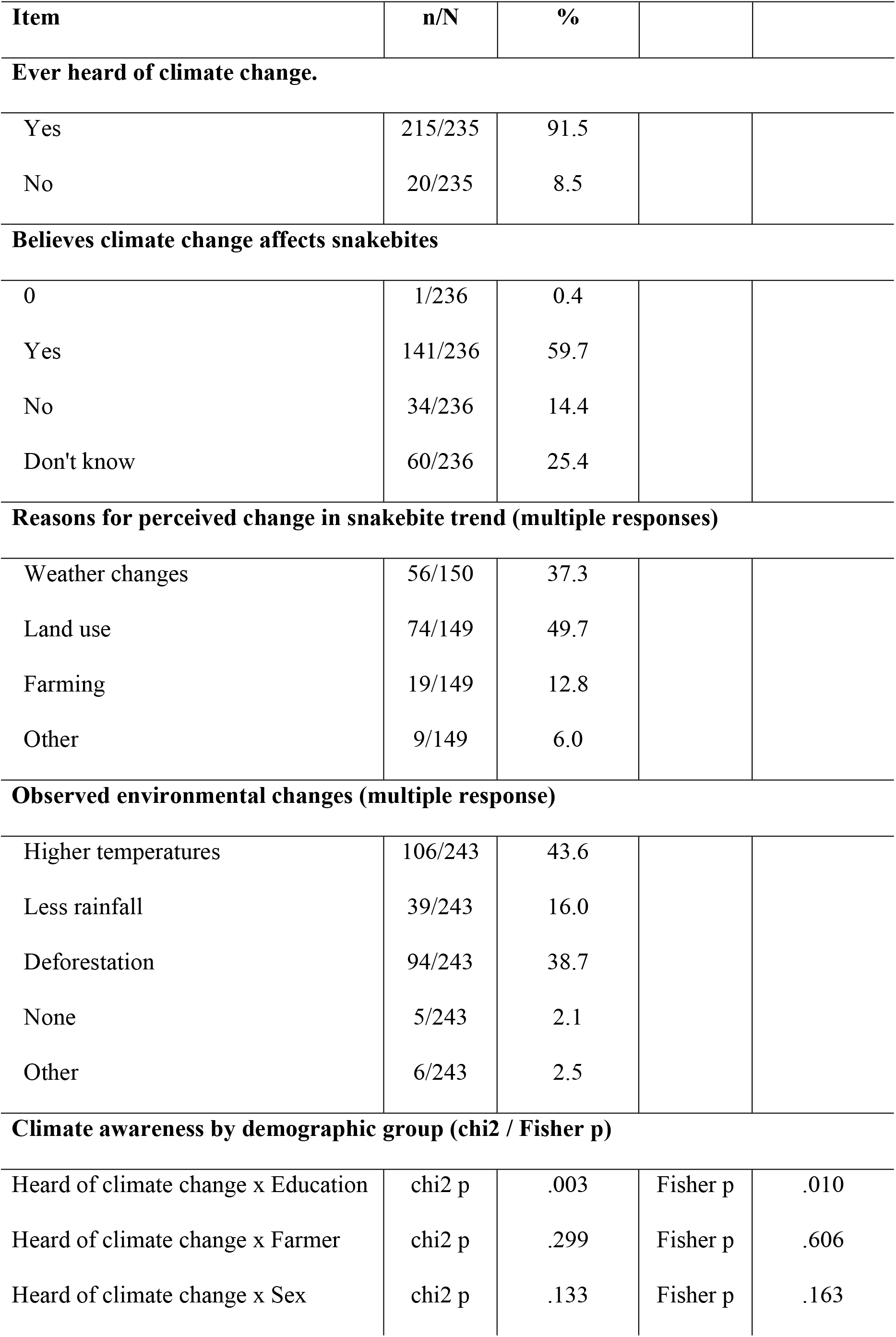
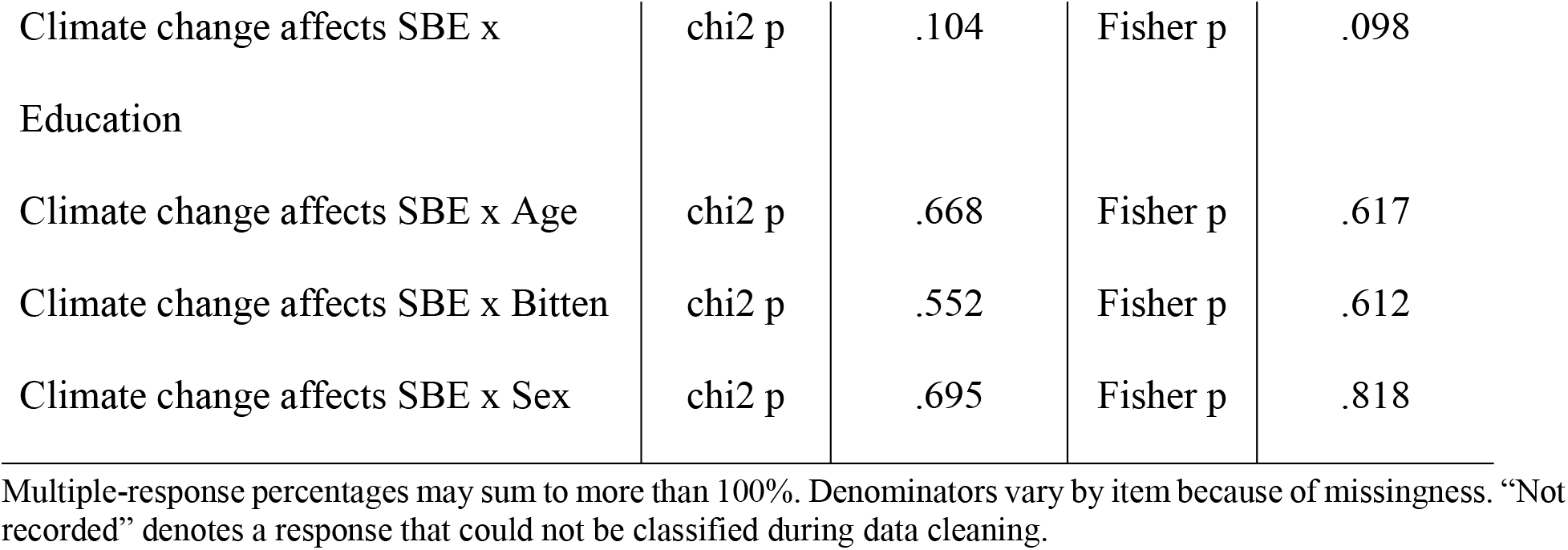
Climate change awareness and perceptions.

| Item | n/N | % |
| --- | --- | --- |

|  |  |  |  |  |
| --- | --- | --- | --- | --- |
| **Ever heard of climate change.** |  |  |  |  |
| Yes | 215/235 | 91.5 |  |  |
| No | 20/235 | 8.5 |  |  |
| **Believes climate change affects snakebites** |  |  |  |  |
| 0 | 1/236 | 0.4 |  |  |
| Yes | 141/236 | 59.7 |  |  |
| No | 34/236 | 14.4 |  |  |
| Don't know | 60/236 | 25.4 |  |  |
| **Reasons for perceived change in snakebite trend (multiple responses)** |  |  |  |  |
| Weather changes | 56/150 | 37.3 |  |  |
| Land use | 74/149 | 49.7 |  |  |
| Farming | 19/149 | 12.8 |  |  |
| Other | 9/149 | 6.0 |  |  |
| **Observed environmental changes (multiple response)** |  |  |  |  |
| Higher temperatures | 106/243 | 43.6 |  |  |
| Less rainfall | 39/243 | 16.0 |  |  |
| Deforestation | 94/243 | 38.7 |  |  |
| None | 5/243 | 2.1 |  |  |
| Other | 6/243 | 2.5 |  |  |
| **Climate awareness by demographic group (chi2 / Fisher p)** |  |  |  |  |
| Heard of climate change x Education | chi2 p | .003 | Fisher p | .010 |
| Heard of climate change x Farmer | chi2 p | .299 | Fisher p | .606 |
| Heard of climate change x Sex | chi2 p | .133 | Fisher p | .163 |

|  |  |  |  |  |
| --- | --- | --- | --- | --- |
| Climate change affects SBE x Education | chi2 p | .104 | Fisher p | .098 |
| Climate change affects SBE x Age | chi2 p | .668 | Fisher p | .617 |
| Climate change affects SBE x Bitten | chi2 p | .552 | Fisher p | .612 |
| Climate change affects SBE x Sex | chi2 p | .695 | Fisher p | .818 |

Multiple-response percentages may sum to more than 100%. Denominators vary by item because of missingness. “Not recorded” denotes a response that could not be classified during data cleaning.

## Discussion

In this rural, agriculture-dependent population in northern Uganda, snakebite emerged as a substantial but largely hidden health and livelihood burden. More than a third of households (36.9%, 95% CI 20.0-57.7) reported at least one lifetime snakebite; among the most recent events, seven of 90 (7.8%) were fatal, and four (4.4%) resulted in permanent disability. More important than the case-fatality proportion, however, was where deaths occurred: five of seven victims died before reaching a health facility. Half of the victims (50.6%) first sought care from a traditional healer, whereas only 2.2% presented initially to a hospital. These findings expose a fundamental limitation of facility-based surveillance. Routine data enumerate people who reach healthcare services, but systematically miss those who die before arrival, remain within informal systems of care, or survive with disability and substantial loss of productive time. The principal contribution of this study is therefore not simply an estimate of snakebite frequency, but evidence that an important component of its burden in rural northern Uganda lies outside the formal health system and is consequently poorly represented in the data used to plan snakebite services [8,9,12]. The predominance of pre-hospital mortality makes the pathway to care a particularly important target for intervention. Palaro lies approximately 50 km from the nearest referral hospital, a distance that possibly contributes both to deaths occurring before arrival and to the under-representation of snakebite fatalities in official health statistics. Traditional healers were the first source of care for approximately half of victims, consistent with reports from elsewhere in sub-Saharan Africa that pluralistic and sequential care-seeking can contribute to delays in definitive treatment [20,21,22]. This pattern should not be interpreted simply as rejection of biomedical care. Most respondents considered hospital treatment effective, and a further third reported that they would use both traditional and formal care. Initial choices appeared instead to reflect proximity, affordability, accessibility and the availability of an immediate local response, factors repeatedly identified as determinants of snakebite care-seeking in rural settings [6,22,23,24,25]. In practice, traditional healers therefore constitute part of the pre-hospital care system, whether or not they are formally recognized as such. Strategies that attempt simply to displace them are likely to have limited effect; a more pragmatic approach would be to engage them in early recognition of severe envenoming, discourage potentially harmful practices such as incision and inappropriate tourniquet use, and establish rapid referral pathways to facilities capable of providing definitive treatment [26]. Such an approach requires prospective evaluation, but it targets the point in the care pathway at which substantial delay appears to arise.

The economic consequences were similarly concentrated outside conventional measures of healthcare expenditure. The median estimated total cost per event was 78,333 UGX (IQR 41,667-160,000), equivalent to approximately 70% of mean monthly household income, despite a median direct monetary cost of zero. Most of the estimated loss arose from foregone productive time. A median of 14 work or school days (IQR 7-21) was lost per event, and permanent disability, reported after 4.4% of the most recent bites, potentially extends this loss indefinitely. This distinction is especially important in a population in which 95% of respondents farmed, and 72.9% reported monthly income below 100,000 UGX. In subsistence agriculture, household labor is both a principal productive asset and a determinant of future food availability; incapacity therefore represents more than lost wages [27,28]. Time lost by victims, together with labor diverted to caregiving, can disrupt cultivation and other livelihood activities even when little or no cash changes hands.

The apparent seasonal concentration of bites adds a potentially important dimension to this economic burden. Reported events clustered in May, coinciding locally with an important period of agricultural activity. Incapacity during such periods could have consequences extending beyond the duration of illness, because delayed planting or other time-sensitive agricultural tasks can affect subsequent production. Our cross-sectional data cannot quantify this downstream effect, and the number of events was insufficient to establish a robust seasonal association. Nevertheless, the convergence of occupational exposure, agricultural timing, and prolonged work loss suggests that snakebite in this setting is appropriately understood as both a health event and a household livelihood shock. Economic evaluations based only on treatment expenditure or other out-of-pocket costs are therefore likely to underestimate its consequences substantially [29,30].

Community perceptions of environmental change provide a further, although more exploratory, dimension to prevention. Awareness of climate change was high (91.5%); 59.7% of respondents believed that climate change affects snakebite risk and a further 25.4% were uncertain. Higher temperatures (43.6%) and deforestation (38.7%) were among the environmental changes most frequently reported, whereas land-use change (49.7%) and weather (37.3%) were commonly reported to explain perceived changes in snakebite patterns. These perceptions are broadly consistent with ecological mechanisms through which temperature, habitat modification, agricultural expansion and changing human-snake contact could alter snakebite risk [13,14,15].

They should not, however, be interpreted as evidence that climate change has caused an increase in snakebite in this population. The cross-sectional design cannot establish temporal changes in incidence, and responses could partly reflect wider exposure to climate change messaging. Their public health relevance lies instead in the degree to which environmental risk is already incorporated into local understandings of snakebite. Apart from an association between education and having heard of climate change (Fisher’s exact p = 0.010), perceptions of a climate-snakebite relationship did not differ substantially by age, sex, education or previous bite experience. Prevention messages could therefore be directed broadly across the community rather than towards a narrowly defined demographic subgroup. More importantly, this study identified seasons, activities and environments in which residents perceived encounters to be more likely. These observations could inform targeted evaluation of practical measures, including protective footwear, lighting, vegetation clearance around homes and modification of high-risk agricultural practices, rather than relying solely on generic snakebite education [14,15,25,28]. Integrating snakebite prevention into existing environmental and climate communication might also provide a culturally intelligible entry point, although the effectiveness of such an approach remains to be tested.

Comparison with other community-based studies suggests that the structure of the problem is regionally consistent, whereas its apparent magnitude requires greater caution. The proportion of victims consulting a traditional healer first (50.6%) was similar to the 53.3% reported across the Kwale and Mbita health and demographic surveillance sites in Kenya [21], somewhat higher than the 42% using traditional treatment either initially or before visiting a facility in the four-county Kenyan household survey by Ooms and colleagues [7], and lower than the 87% reported to have sought informal care first in Eastern Province, Rwanda [31]. These comparisons reinforce the broader observation that formal health facilities frequently represent a later rather than an initial stage in the snakebite care pathway.

By contrast, our estimates of recent snakebite frequency were substantially higher than those reported elsewhere. The five-year household estimate of 29.1% exceeded the corresponding estimates of 5.17% in Kwale and 1.00% in Mbita [21], whereas the 12-month household estimate of 22.3% is difficult to reconcile with published individual-level annual incidence estimates of 4.3 per 1,000 in Eastern Province, Rwanda [31], 665 per 100,000 in Akonolinga, Cameroon [22], 3.7-412.9 per 100,000 across counties in Kenya [32], and approximately 101 per 100,000 previously estimated for Gulu [10]. Direct comparison is complicated by different denominators, case definitions, sampling frames, and the unusually high agricultural exposure of the present study population. These differences are nevertheless unlikely to explain the full magnitude of the discrepancy.

The short-period estimates should therefore be interpreted cautiously. Recall telescoping, in which events occurring outside a specified period are remembered as more recent, provides one plausible explanation and could disproportionately inflate the 12-month and five-year estimates. The lifetime household estimate is less sensitive to temporal displacement and is consequently the more defensible measure of cumulative household exposure in these data, although it too is subject to recall and survivor biases and should not be interpreted as an incidence estimate. Prospective, population-based surveillance with clearly defined person-time denominators will be required to establish contemporary incidence. Importantly, uncertainty over incidence does not negate the principal findings concerning care pathways and consequences: the predominance of pre-hospital deaths, reliance on informal first care, prolonged work loss and permanent disability all indicate forms of burden that facility surveillance is intrinsically poorly positioned to capture.

These findings also complement previous Ugandan evidence on the financial consequences and health-system constraints surrounding snakebite. Catastrophic health expenditure has been reported among 31.4% of affected households in eastern Uganda at the 40% threshold, with the greatest burden among poorer households [29]. Our findings identify a complementary mechanism of economic vulnerability: substantial losses can arise even when direct expenditure is minimal, because illness removes household members from productive activity. Similarly, the pre-hospital deaths observed here provide a community-level counterpart to the treatment delays and limitations in antivenom availability described in Ugandan facility-based studies [12]. Together, these findings suggest that improving antivenom availability, although essential, will be insufficient if patients cannot reach appropriate treatment rapidly.

The policy implication is therefore not a single intervention but a linked pathway of prevention, referral, treatment and recovery. At community level, locally appropriate first-aid education and targeted prevention should address agricultural and household exposures. Traditional healers should be considered potential partners in early recognition and rapid referral, with explicit discouragement of harmful practices. Referral systems need to shorten the interval between bite and definitive care, while appropriate antivenom and trained staff should be reliably available at the lowest level of the health system capable of safely administering treatment. Environmental and climate communication could provide an additional platform for seasonally and occupationally targeted prevention. Finally, because much of the economic burden appears to arise through loss of labor rather than medical expenditure, evaluation of snakebite programs should include disability, time to return to productive activity, caregiver burden and household livelihood effects alongside mortality and antivenom use.

### Strengths and limitations

This study has notable strengths. The sampling frame was probability-based and fully enumerated, and all estimates were survey-weighted to the source population with design-appropriate variance estimation, with the realized design effect reported rather than assumed. To our knowledge, it is one of the very few community-level characterizations of snakebite burden and healthcare-seeking behavior from northern Uganda.

Several limitations should be considered. The 12-month and five-year prevalence estimates were incredibly high relative to the lifetime estimate and to regional data, probably reflecting recall telescoping; we therefore regard the lifetime estimate as the more reliable measure of cumulative household exposure. Snakebites were self-reported and not clinically confirmed, precluding distinction between venomous, non-venomous and dry bites, and all measures were susceptible to recall, misclassification and reporting biases. The cross-sectional design precludes causal inference about seasonality, environmental change, care-seeking or outcomes, while perceived climate-related changes were not independently verified. Multivariable associations were exploratory because clustering reduced the effective sample size and sparse covariate cells limited precision; these findings should therefore be considered hypothesis-generating. Economic estimates based on imputed lost labor should not be interpreted as observed expenditure. Finally, the study was confined to one predominantly agrarian subcounty, limiting generalizability. These limitations constrain estimates of the magnitude and determinants of snakebite, but are less likely to explain the observed pattern of delayed formal care, pre-hospital mortality and livelihood disruption.

## Conclusions

Snakebite in rural northern Uganda is not adequately characterized by the number of patients treated in health facilities. Its burden begins in fields and households, is amplified during the journey to care, and persists through lost labor and disability after the acute clinical episode has ended. Deaths were concentrated before facility arrival, and economic losses were driven predominantly by lost productive labor rather than medical expenditure. Reducing this burden will require earlier access to effective care, engagement of traditional healers in rapid referral, and locally targeted prevention that extends beyond the facility to the communities where exposure occurs, the informal providers whom victims reach first, and the rural households that ultimately absorb the economic consequences of survival.

## Acknowledgments

We thank the participating households and communities of Palaro Subcounty, the research assistants and village health workers, and the local leadership of Gulu District for their support.

## Data availability

A de-identified, cleaned dataset supporting the findings of this study is available in the Zenodo repository (DOI: 10.5281/zenodo.22210575).

## Author contributions

Conceptualisation: FIA. Data curation: FIA, ALI. Formal analysis: FIA. Funding acquisition: FIA. Investigation: FIA, ALI. Methodology: FIA, ELAO, KH, SMN. Project administration: FIA, ALI. Supervision: ELAO, SMN. Writing-original draft: FIA, SHM. Writing-review and editing: KH, SHM, IAB, ELAO, SMN. All authors read and approved the final manuscript.

## Supporting information

**S1. File.** STROBE statement for the study.

**S2. File.** Structured household questionnaire for the study.

